# Age-Stratified SARS-CoV-2 Infection Fatality Rates in New York City estimated from serological data

**DOI:** 10.1101/2020.10.16.20214023

**Authors:** Chloe G. Rickards, A. Marm Kilpatrick

## Abstract

**Importance:** COVID-19 has killed hundreds of thousands of people in the US and >1 million globally. Estimating the age-specific infection fatality rate (IFR) of SARS-CoV-2 for different populations is crucial for assessing the fatality of COVID-19 and for appropriately allocating limited vaccine supplies to minimize mortality.

**Objective:** To estimate IFRs for COVID-19 in New York City and compare them to IFRs from other countries.

**Design, Setting, Participants:** We used data from a published serosurvey of 5946 individuals 18 years or older conducted April 19-28, 2020 with time series of COVID-19 confirmed cases and deaths for five age-classes from the New York City Department of Health and Mental Hygiene. We inferred age-specific IFRs using a Bayesian framework that accounted for the distribution of delay between infection and seroconversion and infection and death.

**Main Outcome and Measure:** Infection fatality rate.

**Results:** We found that IFRs increased approximately 77-fold with age, with a nearly linear increase on a log scale, from 0.07% (0.055%-0.086%) in 18-44 year olds to 5.4% (4.3%-6.3%) in individuals 75 and older. New York City IFRs were higher for 18-44 year olds and 45-64 year olds (0.58%; 0.45%-0.75%) than Spanish, English, and Swiss populations, but IFRs for 75+ year olds were lower than for English populations and similar to Spanish and Swiss populations.

**Conclusions and Relevance:** These results suggest that the age-specific fatality of COVID-19 differs among developed countries and raises questions about factors underlying these differences.

**Key Points:** *Question:* How do age-specific infection fatality rates (IFR) for COVID-19 in the U.S. compare to other populations?

*Findings:* We estimated age-specific IFRs of SARS-CoV-2 using seroprevalence data and deaths in New York City. IFRs increased more than 75-fold with age, from 0.07% in 18-45 year olds to 5.3% in individuals over 75. IFRs in New York City were higher than IFRs in England, Geneva, France and Spain for individuals younger than 64 years old, but similar for older individuals.

*Meaning:* The age-specific fatality of COVID-19 varies significantly among developed nations for unknown reasons.

## Introduction

COVID-19 has killed hundreds of thousands of people in the US and >1 million globally.^1^ The infection fatality rate (IFR) – or the chance of dying after becoming infected – of SARS-CoV-2 is a crucial metric for understanding disease severity. It has been the subject of substantial controversy, in part due to differences between the symptomatic or case fatality rate (CFR) and the infection fatality rate, and also due to differences between naïve estimates of CFR calculated from the ratio of deaths divided by cases and estimates of CFR that account for the delays between infection, case detection and death ^2,3^. Accurately estimating the IFR requires estimates of the number of infections at a point of in time and deaths that result from these infections which occur after a substantial, and highly variable, delay (mean 20.2 d; 95% CI 8.0, 50.0; ^4^). This makes simplistic calculations of IFR problematic when the incidence of infection is changing over time.

Previous analyses have shown enormous variation in IFR with age ^3,5-8^ which indicates that accurate age-specific IFRs are needed to allocate limited supplies of vaccines to meet a goal of minimizing mortality from COVID-19 ^9^. The most accurate approach for estimating age-specific IFRs is a large-scale serosurvey in a population where substantial mortality due to COVID-19 has occurred ^7,8^, because deaths in younger age groups are relatively rare^4^. Substantial differences in age-specific COVID-19 IFRs are apparent in estimates from Spain^8^ and England^7^, indicating the need for additional IFR estimates from other hard-hit populations.

We used data from a seroprevalence study^10^ conducted in New York City in the waning period of the spring epidemic and publicly available death records to estimate age-specific IFRs for the New York city population.

## Methods

We estimated infection fatality rates (IFRs) using data from New York City from a serological survey ^10^ and confirmed COVID-19 cases and deaths recorded by the New York City Department of Health and Mental Hygiene (https://www1.nyc.gov/site/doh/covid/covid-19-data-archive.page). The serosurvey occurred on 6 days over a 10-day period (April 19-28) two-weeks after the peak in COVID-19 cases (Fig 1). Individuals for the serological survey were recruited at grocery stores without prior advertisement to reduce bias and increase randomization ^10^. Daily case counts and number of deaths were obtained from the New York City Department of Health and Mental Hygiene archive webpage for the dates 3/23/20 to 5/17/20 (https://www1.nyc.gov/site/doh/covid/covid-19-data-archive.page).

**Figure 1.**
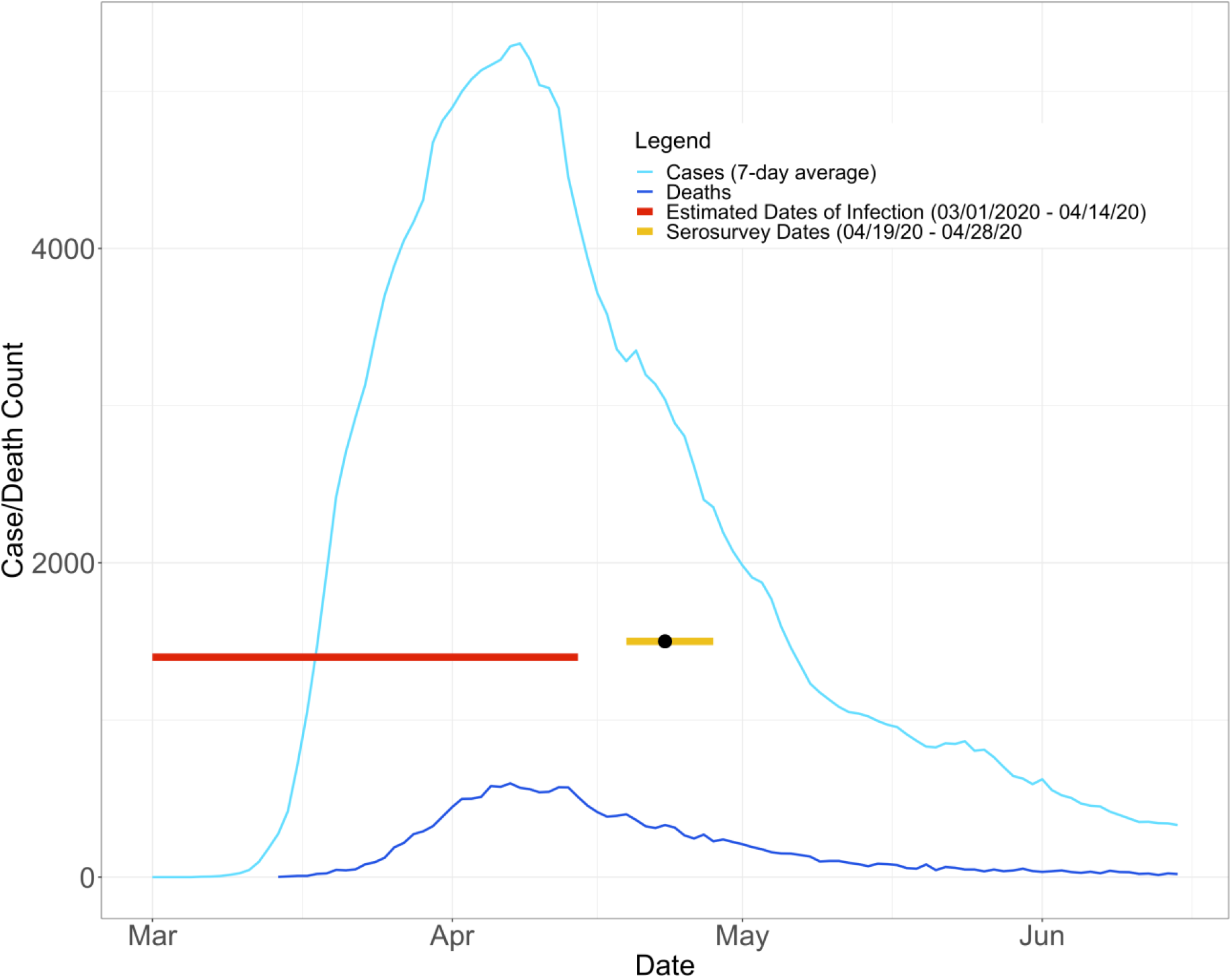
Time series of cases (light blue) and deaths (dark blue) during the spring 2020 COVID-19 outbreak in New York City. The red horizontal segment shows the dates of infections that were likely detected by the midpoint in the 10 day serosurvey, shown by the black point in the yellow segment.

We estimated IFRs using a previously established Bayesian statistical framework ^4^ which combines seroprevalence estimates (including uncertainty) with time series of cases and deaths to estimate the number infected and the fraction dying over time using estimates (log-normal distributions) of the delays between infection, symptom onset, becoming a case, seroconversion, and death ^4^. This approach estimates the cumulative number of infected people at the time of the seroprevalence study and the fraction of those infected that go on to die. We calculated three age-specific estimates of IFRs to address mismatches in age-classes between deaths and seroprevalences and to consider both confirmed and probably COVID-19 deaths (see Supplemental Materials: Methods for details).

We compared the IFR estimates for New York City to two other age-specific estimates based on large serological studies (Paster-Barriuso et al. 2020, Perez-Saez et al. 2020, and Ward et al. 2020), and three IFR estimates based on case fatality rates and near complete testing on closed populations to infer number of infections (cruise ship passengers or repatriated travelers) (Salje et l. 2020, Russell et al. 2020, and Verity et al. 2020). We show IFRs for England and Geneva, Switzerland that include care home resident deaths because the New York City deaths also include care home deaths (IFRs from Spain excluded care home deaths and did not indicate their distribution among age categories). Finally, we show IFRs for both confirmed COVID-19 deaths and excess all-cause deaths for the two studies that included these estimates (Ward et al. 2020 and Pastor-Barriuso et a. 2020).

## Results

The serosurvey in New York City took place in late April, in the latter third of the initial epidemic, when new cases per day had fallen to approximately half of the peak (Fig 1). At this point 22.7% of the population was seropositive, and an estimated 1.5 million infections had occurred ^10^. Due to the delay between infections and deaths, deaths from infections detected in the serosurvey extended into late May (Fig 1). By May 17 there were nearly 16,000 confirmed COVID-19 deaths, and nearly 4,500 probable COVID-19 deaths.

The age-specific IFR for SARS-CoV-2 in New York City increased with age approximately 77-fold from 0.07% in 18-44 to 5.3% in 75+ year-olds, in the raw analysis where we assumed equal seroprevalence for all subgroup age-classes (55-64, 65-74, 75+) within the 55+ age-class (Table 1). In the adjusted analysis where used fine-scale age-specific seroprevalence data from Spain to estimate seroprevalence of 0-17 year-olds and variation within the 55+ age class, the IFR increased from 0.0018% for 0-17 year-olds, or roughly 2 fatalities per 100,000 infected individuals to 5.4% in 75+ year-olds (Table 1). Including probable COVID-19 deaths increased IFRs 20-31% across the five age classes (Table S1).

**Table 1.**
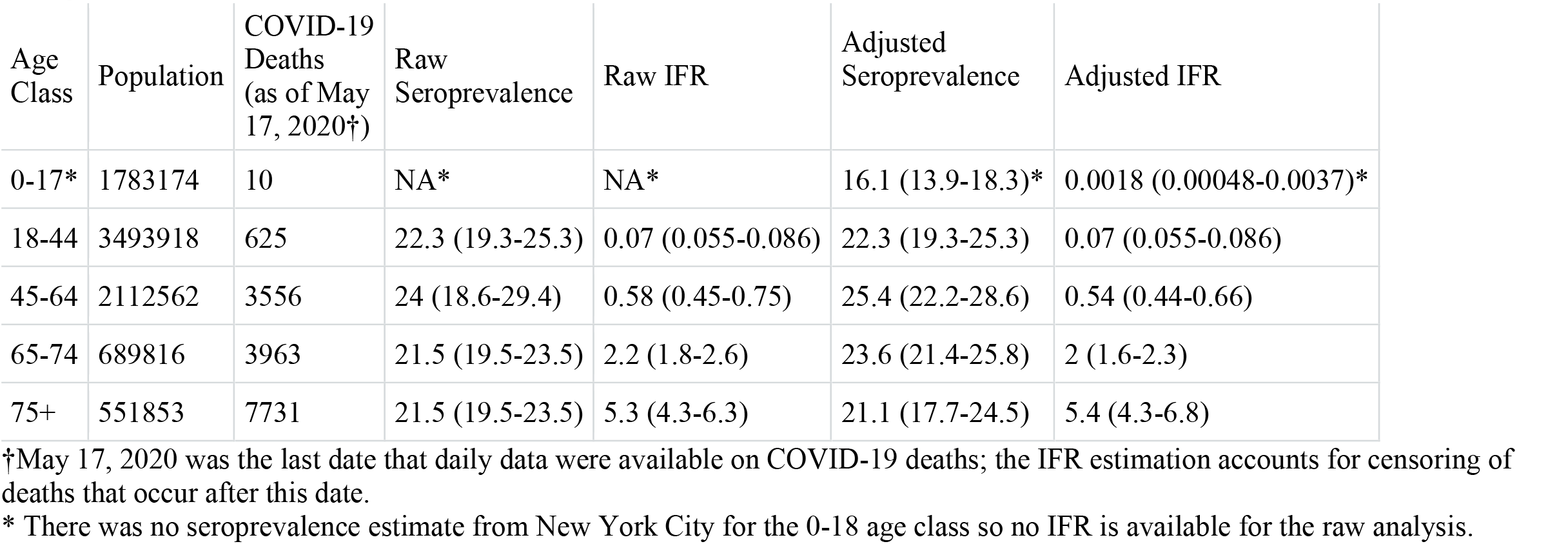
Seroprevalence and infection fatality rate (IFR) estimates in New York City for five age classes using the raw values ^10^ or adjusted values (See Methods for details).

| Age Class | Population | COVID-19 Deaths (as of May 17, 2020†) | Raw Seroprevalence | Raw IFR | Adjusted Seroprevalence | Adjusted IFR |
| --- | --- | --- | --- | --- | --- | --- |
| 0-17* | 1783174 | 10 | NA* | NA* | 16.1 (13.9-18.3)* | 0.0018 (0.00048-0.0037)* |
| 18-44 | 3493918 | 625 | 22.3 (19.3-25.3) | 0.07 (0.055-0.086) | 22.3 (19.3-25.3) | 0.07 (0.055-0.086) |
| 45-64 | 2112562 | 3556 | 24 (18.6-29.4) | 0.58 (0.45-0.75) | 25.4 (22.2-28.6) | 0.54 (0.44-0.66) |
| 65-74 | 689816 | 3963 | 21.5 (19.5-23.5) | 2.2 (1.8-2.6) | 23.6 (21.4-25.8) | 2 (1.6-2.3) |
| 75+ | 551853 | 7731 | 21.5 (19.5-23.5) | 5.3 (4.3-6.3) | 21.1 (17.7-24.5) | 5.4 (4.3-6.8) |

†May 17, 2020 was the last date that daily data were available on COVID-19 deaths; the IFR estimation accounts for censoring of deaths that occur after this date.
\* There was no seroprevalence estimate from New York City for the 0-18 age class so no IFR is available for the raw analysis.

The age-specific IFRs from New York City for the 18-44 and 45-64 age classes were higher (with non-overlapping 95% confidence intervals) than corresponding IFRs for England, Geneva, France and Spain, but had overlapping confidence intervals and were similar to estimates for China (Figs. 2, S2). In contrast, the IFRs for the oldest two age classes, 65-74 and 75+ were lower (with non-overlapping 95% CIs) than IFRs from England, but had overlapping confidence intervals and were similar to corresponding IFRs from Geneva, France, Spain and China (Figs 2, S2).

**Figure 2.**
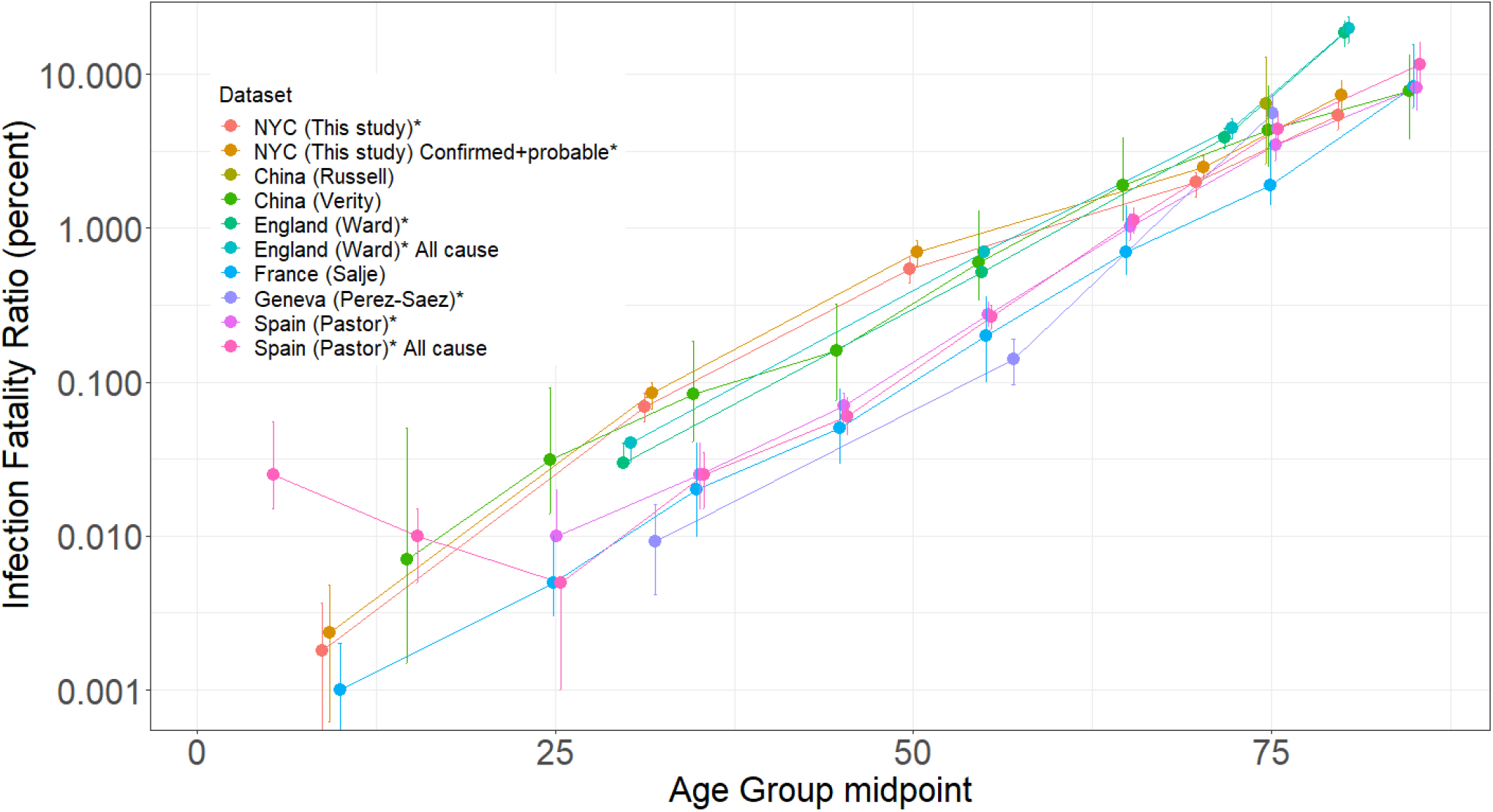
Age-specific infection fatality ratio (IFR) of COVID-19 on a log-scale (mean ± 95% CI) with points plotted at the midpoint of the age-class on the x-axis (points are slightly jittered along the x-axis facilitate presentation). Lines show age-specific IFRs for different populations, either including just confirmed COVID-19 deaths, or estimates from “All cause” mortality. For New York City (This study) the figure shows the “adjusted” estimates from Tables 1 and S1 for both confirmed COVID-19 deaths and confirmed + probable COVID-19 deaths. Datasets with an asterisk (*) estimate the number of infections based on seroprevalence studies; others estimate infections using either testing of passengers on the Princess Cruise ship (Russell, Salje) or repatriated travelers from Wuhan (Verity).

## Discussion

The IFRs in New York City were higher for the two younger age classes (18-44, 45-64), which accounted for 26% of the 15,885 deaths (Table 1), than all three other studies based on large-scale serosurveys (Geneva, Spain and England). This may have been due to a higher prevalence of pre-existing conditions in these age-groups than in other populations; 79% and 85% of COVID-19 deaths in these two age groups had pre-existing conditions (Diabetes, Lung Disease, Cancer, Immunodeficiency, Heart Disease, Hypertension, Asthma, Kidney Disease, GI/Liver Disease, or Obesity). In contrast, IFRs for the two older age classes were very similar to estimates from most other countries despite 79% and 76% of these deaths also having pre-existing conditions.

Two other factors that influenced both IFRs or exposure to SARS-CoV-2 in previous studies are sex and race/ethnicity ^7,8,10^. Although neither seroprevalence nor daily age-specific COVID-19 deaths by sex and race/ethnicity were publicly available, crude calculations can be made assuming that differences in exposure and death between ages were consistent among sexes and races/ethnicities. There were 55% more deaths in New York City in men than women, whereas seroprevalence was only 6.3% higher in men than women ^10^, suggesting that IFRs for men were, on average across ages, 46% lower. This is a smaller sex difference than estimated for Spanish and French populations ^6,8^. Seroprevalence in New York City was 1.98-fold higher for hispanic/latinos than white people^10^ whereas death rates were 5.9, 3.1, 2.5, and 1.6-fold higher for 18-44, 45-64, 65-74, and 75+ age-classes, respectively, suggesting that either age-specific exposure or age-specific IFRs differ among these races/ethnicities.

Two other studies have estimated IFRs for New York City using other approaches. Our age-specific estimates for IFRs in New York City were uniformly lower (by 1.7 to 2.6-fold) than IFRs estimated from model-based inference of infections ^11^, and crudely similar to, but difficult to compare to, a crude ratio of deaths four weeks after a serosurvey divided by estimated infections from that serosurvey, because that study^5^ estimated IFR for age categories than that didn’t match the published seroprevalence study^10^ or health department death data.

One shortcoming of our study is that the oldest age class for the seroprevalence study was broad (55+) and combined ages (60-69, 70-79, 80+) in which IFRs differed 10-fold in other studies ^8^, whereas deaths were available in finer age classes. The two analyses we performed to quantify IFRs within this 55+ group produced similar results because the fine resolution seroprevalence data from Spain suggested only a slightly lower seroprevalence for 75+ compared to 45-54 (∼4.4%; Table S2). In contrast, seroprevalence in England for 65-74 and 75+ age classes was 1.8 times lower than for 55-64 year-olds (Fig S3); a non-representative serosurvey of residual sera from commercial labs^12^ also showed lower seroprevalence in older age classes. If we assume the relative seroprevalence in New York City in these three age classes was similar to England, IFRs for the latter two groups would be 1.8-fold higher: 3.6%, and 9.5%. This IFR for the oldest group is still substantially lower than the corresponding IFR for England (Fig 2, S2). The variation in seroprevalence within older age classes in some studies^7,13^ emphasizes the importance of reporting both seroprevalence and deaths for SARS-CoV-2 for finer age classes to enable a more accurate understanding of the fatality of COVID-19 for these older age groups which are most vulnerable to death from COVID-19. A key unanswered question concerning the fatality of COVID-19 is how much age-specific IFRs have changed over time due to better case management and treatments.

## Data Availability

All data in the ms are publicly available and links are provided to these sources.

## Acknowledgements

Funding was provided by NSF grant DEB-1911853. We thank Javier Perez-Saez and co-authors for making their R code available (https://github.com/HopkinsIDD/sarscov2-ifr-gva) which facilitated the analyses presented here.

## Supplemental Tables and Figures

**Table S1.** Seroprevalence and infection fatality rate (IFR) estimates for five age classes using both confirmed and probable COVID-19 deaths and either the raw seroprevalence values in ^10^ or adjusted values (See methods for details).

| Age Class | Population | Deaths | Raw Seroprevalence | Raw IFR | Adjusted Seroprevalence | Adjusted IFR |
| --- | --- | --- | --- | --- | --- | --- |
| 0-17 * | 1783173.8 | 13 | NA* | NA* | 16.1 (13.9-18.3) | 0.0031 (0.0013-0.0058) |
| 18-44 | 3493918.2 | 750 | 22.3 (19.3-25.3) | 0.085 (0.068-0.1) | 22.4 (19.4-25.4) | 0.085 (0.067-0.1) |
| 45-64 | 2112562 | 4490 | 24 (18.6-29.4) | 0.75 (0.57-0.95) | 25.4 (22.4-28.4) | 0.7 (0.57-0.83) |
| 65-74 | 689816 | 4893 | 21.5 (19.5-23.5) | 2.8 (2.3-3.3) | 23.6 (21.4-25.8) | 2.5 (2.1-3) |
| 75+ | 551853 | 10105 | 21.5 (19.5-23.5) | 7.1 (5.8-8.4) | 21 (17.6-24.4) | 7.3 (5.8-9.1) |

\* There was no seroprevalence estimate for the 0-17 age class so no IFR is available for the raw analysis.

**Fig S1.**
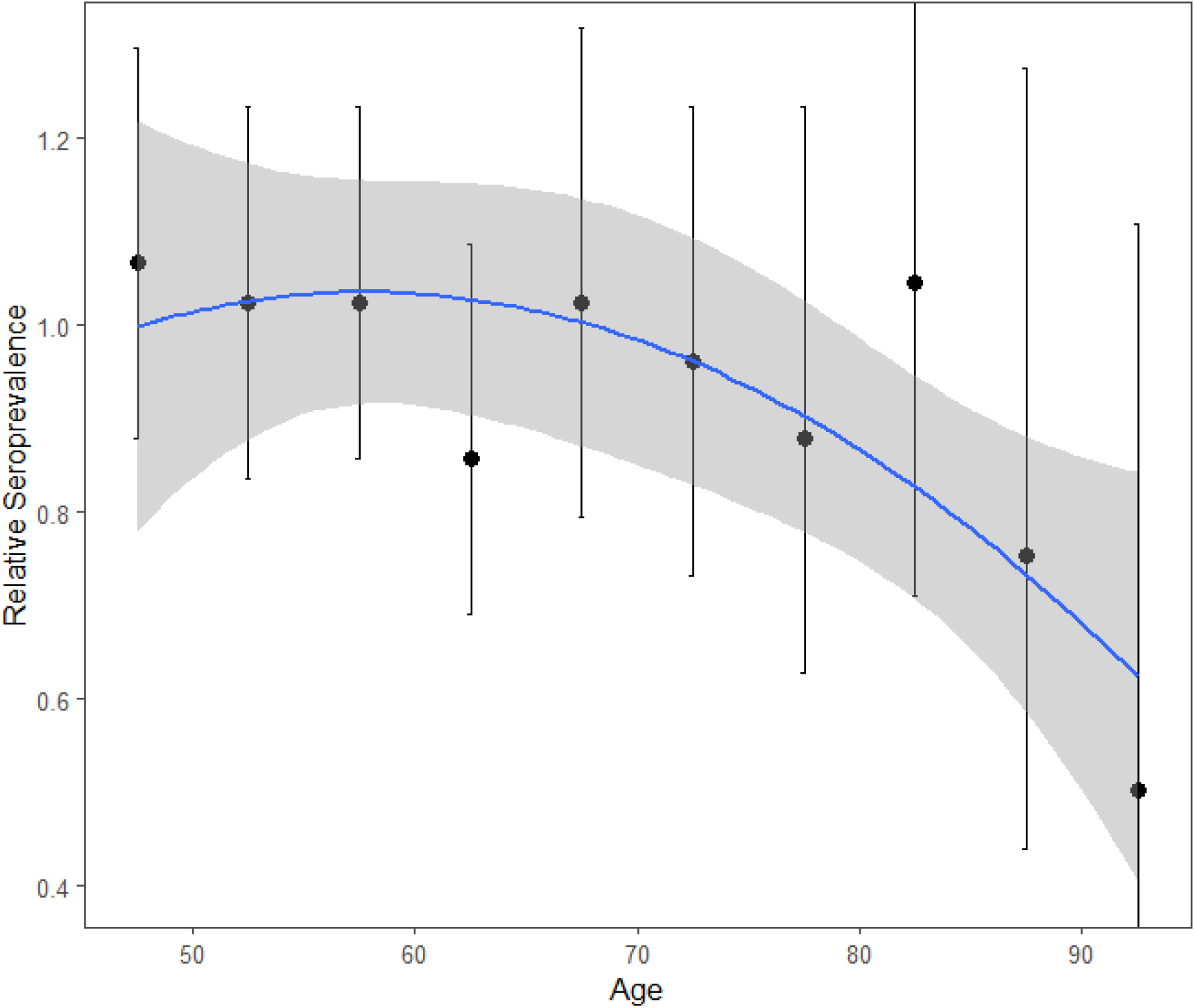
Variation in relative SARS-CoV-2 seroprevalence in Spain ^13^ versus age among those 45 years and older (see Fig S3 for absolute seroprevalence values for all ages). Points show seroprevalence divided by the mean seroprevalence of individuals younger than 70 and curve shows quadratic fit to points (R^2^ = 65%).

**Table S2.**
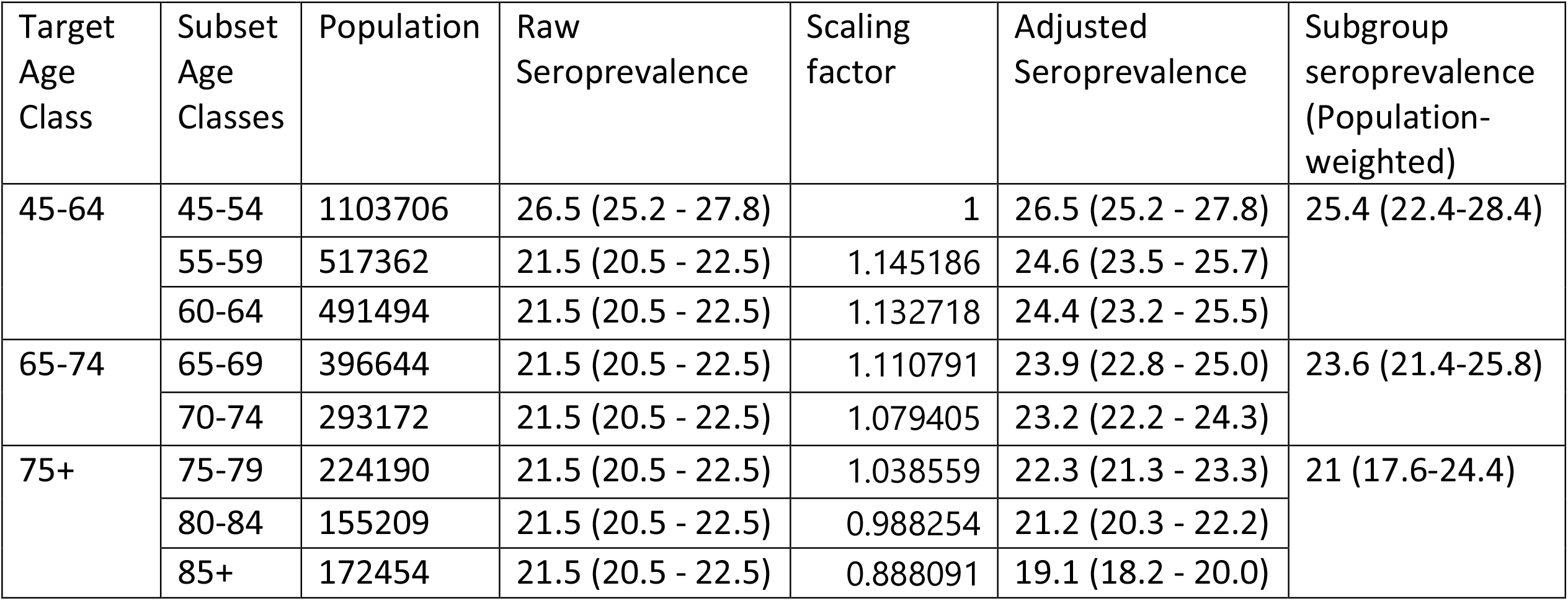
Detailed calculations for estimating the subgroup seroprevalence for the 55+ age class into seroprevalence estimates for the 45-65, 65-75, and 75+ age classes using the relative seroprevalence values shown in Figure S1.

| Target Age Class | Subset Age Classes | Population | Raw Seroprevalence | Scaling factor | Adjusted Seroprevalence | Subgroup seroprevalence (Population-weighted) |
| --- | --- | --- | --- | --- | --- | --- |
| 45-64 | 45-54 | 1103706 | 26.5 (25.2 - 27.8) | 1 | 26.5 (25.2 - 27.8) | 25.4 (22.4-28.4) |
|  | 55-59 | 517362 | 21.5 (20.5 - 22.5) | 1.145186 | 24.6 (23.5 - 25.7) |  |
|  | 60-64 | 491494 | 21.5 (20.5 - 22.5) | 1.132718 | 24.4 (23.2 - 25.5) |  |
| 65-74 | 65-69 | 396644 | 21.5 (20.5 - 22.5) | 1.110791 | 23.9 (22.8 - 25.0) | 23.6 (21.4-25.8) |
|  | 70-74 | 293172 | 21.5 (20.5 - 22.5) | 1.079405 | 23.2 (22.2 - 24.3) |  |
| 75+ | 75-79 | 224190 | 21.5 (20.5 - 22.5) | 1.038559 | 22.3 (21.3 - 23.3) | 21 (17.6-24.4) |
|  | 80-84 | 155209 | 21.5 (20.5 - 22.5) | 0.988254 | 21.2 (20.3 - 22.2) |  |
|  | 85+ | 172454 | 21.5 (20.5 - 22.5) | 0.888091 | 19.1 (18.2 - 20.0) |  |

**Figure S2.**
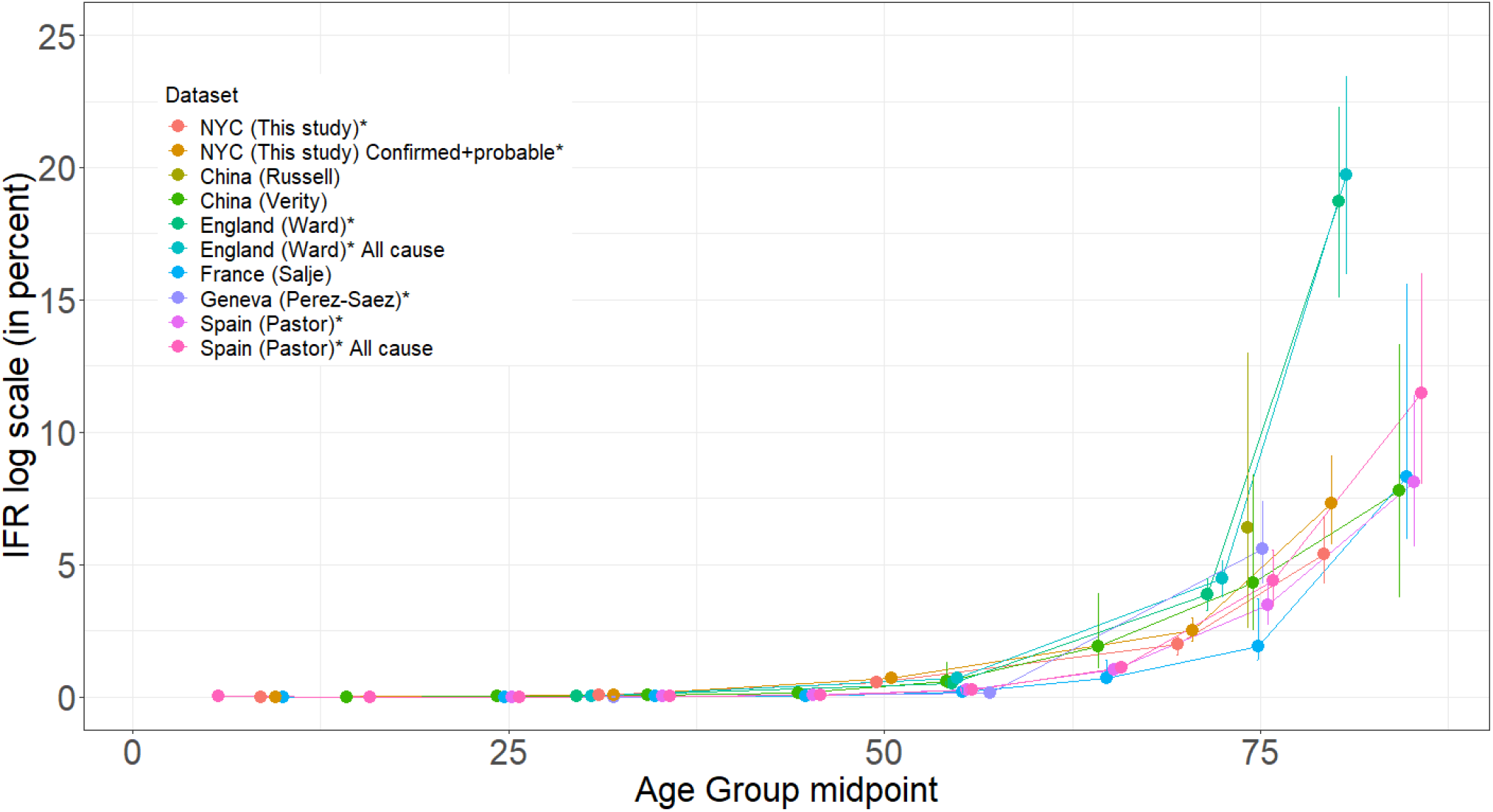
Age-specific infection fatality ratio (IFR) of COVID-19 (mean ± 95% CI) on an untransformed scale with points plotted at the midpoint of the age-class on the x-axis (points are slightly jittered along the x-axis facilitate presentation). Lines show age-specific IFRs for different populations, either including just confirmed COVID-19 deaths, or estimates from “All cause” mortality. For New York City, “NYC (This study)” the figure shows the “adjusted” estimates from Tables 1 and S1 for both confirmed COVID-19 deaths and confirmed + probable COVID-19 deaths. Datasets with an asterisk (*) estimate the number of infections based on seroprevalence studies; others estimate infections using either testing of passengers on the Princess Cruise ship (Chia (Russell), France (Salje)) or repatriated travelers from Wuhan (China(Verity)).

**Fig S3.**
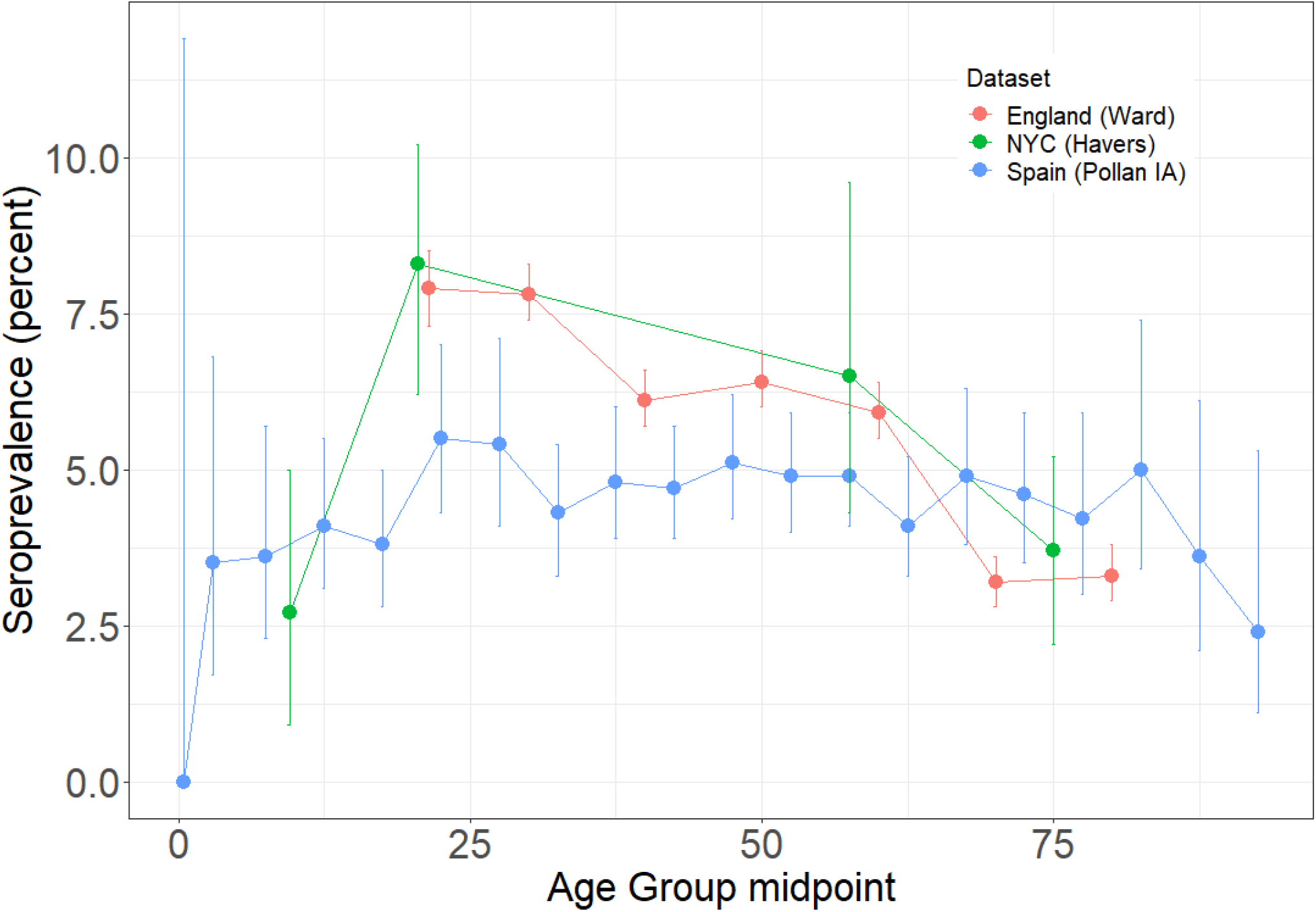
Variation in seroprevalence by age (mean ± 95% CI) from two large representative serosurveys (Spain; data shown for the immunoassay, IA; and England) and one serosurvey using residual sera from commercial laboratories (New York City, NYC).

## Supplemental Methods

The age classes used for the seroprevalence study^10^ (18-34, 35-44, 45-54, and 55+) did not match the age classes of the cases and deaths provided by the New York City Department of Health (0-17, 18-44, 45-64, 65-74, and 75+). Most importantly, the seroprevalence study grouped all individuals older than 55 and did not include individuals younger than 18. Previous estimates of IFR differ 30-fold between 50-59 (0.38%; 95% CI: 0.32-0.45), and 80+ (11.62%; 95% CI: 8.06–16.47) ^8^, so we estimated IFRs for subgroups within the 55+ group by using the finer age classes reported by the New York City Department of Health for cases and deaths (55-64, 65-74, and 75+). As a result, we performed two separate analyses using different estimates of seroprevalence for these finer age classes.

For the first (“raw”) analysis, we used the same single seroprevalence estimate from the 55+ group for all three age classes that we split this group into: 55-64, 65-74, and 75+. For this raw analysis we did not estimate the IFR for the 0-17 age class. We estimated the seroprevalence in each of the age-classes for the death data using population-weighted averages from the seroprevalence study. We used 2017 estimates of 2010 census data for population estimates (https://www.baruch.cuny.edu/nycdata/population-geography/age_distribution.htm). For example, for the IFR for the 45-64 age class, we averaged the seroprevalence estimates for the 45-54, and 55+ age classes, weighted using the fraction of the New York City population that are in the 45-54 and 55-64 age classes.

For the second (“adjusted”) analysis, we used fine-resolution (5-year increments, from infancy to 85+) seroprevalence data from a study in Spain ^13^ (Fig. S3), to estimate a seroprevalence for the 0-18 group in which 10 deaths due to SARS-CoV-2 occurred in New York City in this age group and to estimate variation in seroprevalence with the 55+ age group for the three oldest age classes 55-64, 65-74, and 75+. We fit a simple quadratic function to the data, which captured the decrease with age in seroprevalence after age 55 (Fig S1). We calculated population-weighted estimates of prevalence for the subgroups within the 55+ age class, which resulted in elevated seroprevalence in 55-64 age class and lower seroprevalence in the 65-74 and 75+ age classes relative to the 55+ age class (Table S1). We also used data for individuals 0-17 from Spain to estimate seroprevalence for this age group in New York City. The ratio of seroprevalence in 0-17 year olds to seroprevalence in 18-44 year olds in Spain ^13^ was 0.72. We multiplied the seroprevalence in the 18-44 age class from the study in New York City by this relative ratio to estimate seroprevalence of 0-17 year olds (Table 1).

